# Precision diagnosis for monogenic diabetes requires ethnicity specific criteria for genetic testing

**DOI:** 10.64898/2026.02.05.26345659

**Authors:** Sophie Jones, Julianne Knupp, Snehal Pandya, Olivia Groom, Cleo Goodall, Ann Sebastian, Kevin Baynes, Srikanth Bellary, Anna Brackenridge, Mohammed SB Huda, Rajni Mahto, Jayanti J Rangasami, Shenaz Ramtoola, Andrew T Hattersley, Des G Johnston, Kevin Colclough, Beverley Shields, Jayne AL Houghton, Shivani Misra

## Abstract

The detection of monogenic diabetes illustrates the potential of precision medicine, with treatments tailored to specific genes and diagnosis involving targeted genetic testing. Current detection criteria are derived from White populations. We investigated detection of monogenic diabetes in an unselected multiethnic cohort comprising 1,706 participants diagnosed with diabetes before the age of 30-years.

Using broad biomarker criteria (triple pancreatic antibody negative and detectable C-peptide) to select for next generation sequencing of monogenic diabetes genes, we found a non-significantly different minimum cohort prevalence of monogenic diabetes of 2.1% in White, 2.0% in South Asian, 2.5% in African-Caribbean, and 3.6% in Mixed participants. The detection rate, however, varied significantly (17.7% in White, 5.3%in South Asian, 8.0% in African-Caribbean, and 15.2% in Mixed participants, p<0.001). Those without monogenic diabetes showed significant variations in BMI. No difference in phenotype of monogenic diabetes across ancestry groups was observed. Non-white ethnicity participants were significantly more likely to have undiagnosed monogenic diabetes than White with on average a 10-year duration before receiving a correct diagnosis. By applying ancestry-specific BMI cut-offs (White <30, South Asian <27, African-Caribbean and Mixed <35 kg/m²), the overall detection rate increased from 8.8 to 16%, reducing the number needed to test to identify one case from 11 to 6 and boosting detection rates to 39, 11, 9 and 26% in White, South Asian, African-Caribbean and Mixed-ethnicity participants, respectively. These findings were validated in an external real-world dataset.

Applying broad biomarker criteria for initial selection, mitigates clinical biases leading to misclassification of monogenic diabetes in non-White ethnicities. However, further tailoring criteria with ethnic-specific BMI cut-offs doubled detection rates, improving cost-effectiveness by minimising unnecessary testing. Our study highlights the need to develop precision medicine approaches accounting for phenotypic variation across diverse populations, to ensure accurate diagnoses and cost-efficient healthcare provision.

## Introduction

Maturity onset diabetes of the young (MODY), the most common form of monogenic diabetes, accounts for 1-3% of adults with diabetes from White European ethnicity^1,2^. Treatment depends on the affected gene and individuals achieve superior glycaemic control on gene-specific treatment, rather than the standard care for type 1 or type 2 diabetes^3^.

The detection and treatment of monogenic diabetes represents a major advance in the delivery of precision medicine in diabetes, however systematic studies to identify the best criteria for detection of monogenic diabetes, have predominantly been undertaken in White European populations^4^. Current guidelines and consensus recommendations are therefore based on data derived from the study of those of White European ancestry^5^.

Identifying monogenic diabetes amongst other types is clinically challenging. In a UK study, 80% of people with MODY were estimated to be misdiagnosed with type 1 or type 2 diabetes^6^. Routine genetic testing for all patients with early-onset diabetes is not feasible due to associated costs. Diagnostic approaches have therefore prioritised stratifying patients based on key clinical features for targeted molecular genetic testing. This method aims to maximise case identification while minimising unnecessary testing in the broader population. Current algorithms for the detection of MODY centre on the use of clinical data such as age-at-diagnosis, parental history and treatment along with biomarkers, such as C-peptide and pancreatic autoantibodies, which are used to distinguish MODY from type 1 diabetes^1^. Such approaches alone or combined in a MODY probability calculator have been shown to identify a subset of individuals most likely to have monogenic diabetes in White individuals, with almost a 20-30% detection rate^1^.

The detection of monogenic diabetes in non-White European populations is less well studied.

In two previous studies of centralised national genetic testing laboratories examining detection rates for MODY, the detection in UK South Asian individuals^7^ and in ‘non-white’ individuals grouped together^8^ (predominantly North African) was significantly lower than White European individuals.

Using patient data from genetic testing registries to establish optimal ethnicity-specific criteria introduces bias due to the lack of standardisation in the reasons for referral and the selection criteria applied by clinicians, who are likely to pre-select patients with a higher likelihood of having MODY.

In the few unselected studies of children and adolescents’ monogenic diabetes cases were found in all ancestry groups studied, but the characteristics by ancestry groups were not reported^9,10^.

In the present study we aimed to address these knowledge gaps using data from a cross-sectional study of early-onset diabetes with the aim of 1) ascertaining the detection rate of monogenic diabetes across White, South Asian and African or Caribbean UK participants using unbiased selection criteria and 2) determining the optimal referral criteria for genetic testing by ancestry.

## Methods

### Recruitment

The MY DIABETES Study (clinicaltrials.gov NCT02082132) was an observational cross-sectional study recruiting individuals from White European, South Asian and African or Caribbean ancestry and a diagnosis of any type of diabetes under the age of 30-years, with any current-age. The study received ethical approval from the Chelsea Local Research Ethics Committee (13/LO/0944). Recruitment took place from hospital-based adult and paediatric diabetes clinics at 41 hospitals across England from 2014 - 2021 targeting centres located in areas with ethnic diversity. Exclusion criteria included an established secondary cause of diabetes or another ethnic group. People with mixed ethnicity were also recruited.

### Study Visit

Participants attended a single study visit at their local centre and provided written informed-consent. Where possible, individuals were asked to attend fasting, however children were not required to fast.

The 1-hr study visit comprised a detailed clinical history (diabetes diagnosis, symptoms, treatments and complications, family history, self, parental and grand-parental ancestry), anthropometry and blood tests, including glycated haemoglobin (HbA1c), lipid profile, C-peptide, pancreatic auto-antibodies and whole blood for DNA extraction. For the first year of the study anti-Glutamic Acid Decarboxylase (GAD-65), and anti-Islet Antigen-2 (IA2) antibodies were measured, and from the second year of the study, anti-zinc transporter 8 (ZnT8) measurement was additionally undertaken.

### Data Handling

Age-at-diagnosis of diabetes was self-reported by participants. Sex at birth was recorded. During analysis of family history, adopted and half siblings were excluded. For analysis, ethnicity was categorised as White European, South Asian, African or Caribbean, Mixed or unknown for each individual. South Asian was defined as ancestry from India, Bangladesh, Sri Lanka or Pakistan. In all cases, if the self-reported ethnicity and that of all four grandparents matched, an individual was categorised as being from that ethnic group. If the ethnicity of two grandparents was unknown, but that of the corresponding parent was known and matched self-reported ethnicity and that of the other grandparents, a participant was also classed to be from that group. Where a participant’s grandparents belonged to different ethnicity categories, they were classed as Mixed. Participants with unknown ethnicity or those with ethnic groups other than that specified above were excluded from subsequent analyses. BMI analyses were restricted to those >18 years at the time of study.

### Genetic Testing Pathway

Those with at least one positive autoantibody or undetectable C-peptide (<27 pmol/L) were classified as having type 1 diabetes and excluded from genetic testing. Participants negative for anti-GAD-65, anti-IA2, and anti-ZnT8 antibodies, with C-peptide _≥_27 pmol/L, underwent genetic testing for monogenic diabetes [broad biomarker criteria]. Antibodies were assayed using RSR Ltd immunoassays, with positivity defined (according to 97^th^ centile) as _≥_11 U/mL (GAD), _≥_7.5 U/mL (IA-2), and _≥_10 U/mL or _≥_65 U/mL (ZnT8 for adults <30 or >30 years). C-peptide was analysed using the Abbott Architect platform (lower detection limit: 27 pmol/L).

### Genetic Analysis

Genetic analysis to identify monogenic diabetes cases was performed using a targeted next generation sequencing panel of 54 genes and 19 mitochondrial DNA variants associated with MODY and maternally inherited mitochondrial diabetes. This was performed at the Exeter Genomics Laboratory at Royal Devon University Healthcare NHS Foundation Trust, Exeter, UK. All probands identified in this study had variants classified as likely pathogenic (class 4) or pathogenic (class 5) based on established guidelines^11^. Participants with variants of unknown significance were not included in the final analysis.

### Applying additional criteria to optimise detection

After completion of genetic testing, we retrospectively applied additional selection parameters to the participants who had undergone genetic testing to test diagnostic accuracy of additional parameters. We tested the effectiveness of additional BMI thresholds applied to the broad biomarker criteria <35, <30, <27.5 kg/m^2^ across each ethnic group and identified the best criteria for each group to ‘optimise’ the overall detection rate. We applied these BMI thresholds in both Adults and Children for comparability, recognising that BMI has some value in identifying obesity even in children^12^.

### Outcomes

Cohort prevalence was determined as the percentage of probands with confirmed pathogenic mutations within each ethnic group expressed as a percentage of all participants recruited. Detection rate (positive predictive value) was calculated as the percentage of probands with confirmed mutations among those meeting specific criteria for genetic testing. False negative rate was defined as the proportion of MODY cases missed by each criterion (expressed as a proportion of the total number of MODY cases that could have been found). Additionally, we calculated negative predictive value and the number needed to test to identify one case. All analyses were stratified by ethnicity.

### External Validation

We applied the BMI cut-offs defined in the above analysis to the monogenic diabetes genetic testing referral database held at the Exeter Molecular Genetics Laboratory, England. This registry includes a diverse cohort of patients enriched for monogenic diabetes who have been referred for genetic testing by their clinician. We examined detection rates in all referrals that underwent genetic testing by BMI, stratified by ethnic group (restricted to White, Asian and Black). We calculated numbers needed to test at specific BMI thresholds.

We further supplemented this real-world validation of our proposed cut-offs with population-level data to ascertain proportions of individuals would be eligible for genetic testing by ethnicity according to proposed BMI cut offs. Using Northwest London electronic healthcare record data, accessed via Whole Systems Integrated Care database, we extracted individuals coded with type 2 diabetes between 1^st^ January 2010 and 24^th^ January 2026. We included those with a recorded diagnosis <30 years and those recorded as White, South Asian or Black ethnicity. We excluded individuals with other types of diabetes codes and individuals who had not reached 18 years of age by end of follow-up were excluded in order to study BMI. Using this cohort, we extracted the latest BMI on record and analysed proportions eligible for testing in each ethnic group, by BMI threshold.

### Statistical Analysis

Descriptive statistics are presented as n (percentage) for categorical variables and median (interquartile range, IQR) for non-parametric continuous variables. Comparisons across ethnic groups used a two-way chi-squared test for categorical variables and Mann-Whitney U or Kruskal-Wallis tests for continuous variables. Statistical significance was set at <0.05 and all analyses were performed using STATA version 18.

## Results

### Cohort Demographics

Of 1,785 participants recruited, 1,706 met criteria for analysis (figure 1), comprising 653 White, 639 South Asian, 275 African Caribbean and 139 people with Mixed ethnicity (table 1). Overall n=925 (54%) were females with no significant difference in gender distribution by ethnicity (p=0.31). The median age at diagnosis was 16.0 years (10.0 – 23.9) with a duration of 13.8 (5.8 - 24.7) years. Current age and duration varied significantly by ethnic group; age at recruitment was oldest in White 36.0 years (27.1-50.2) and lower in all other ethnic groups (p<0.001), table 1.

**Figure 1:**
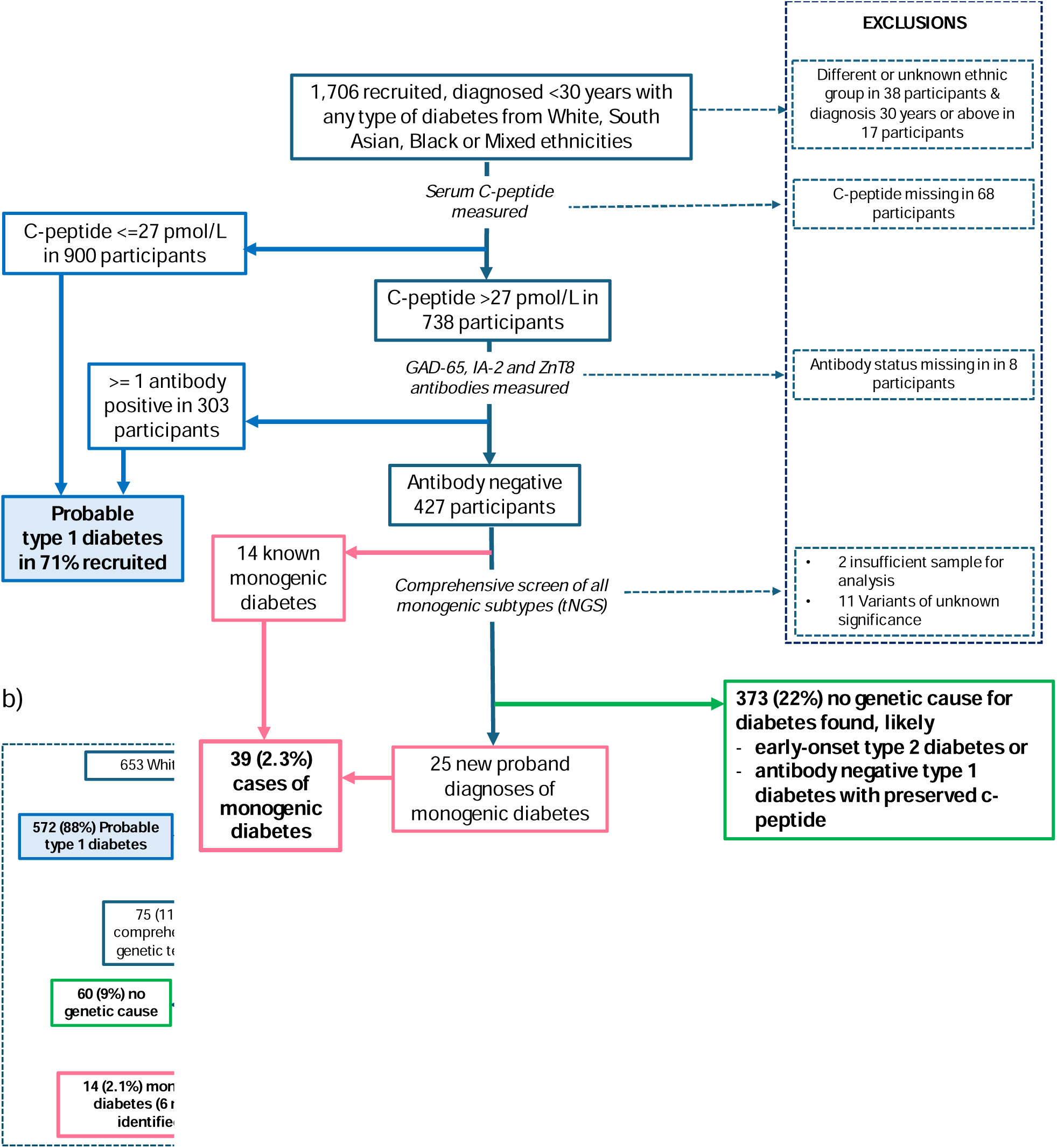
Flow chart depicting the biomarker pathway and end diagnoses of a) all participants and b) by each ethnic group

**Table 1:**
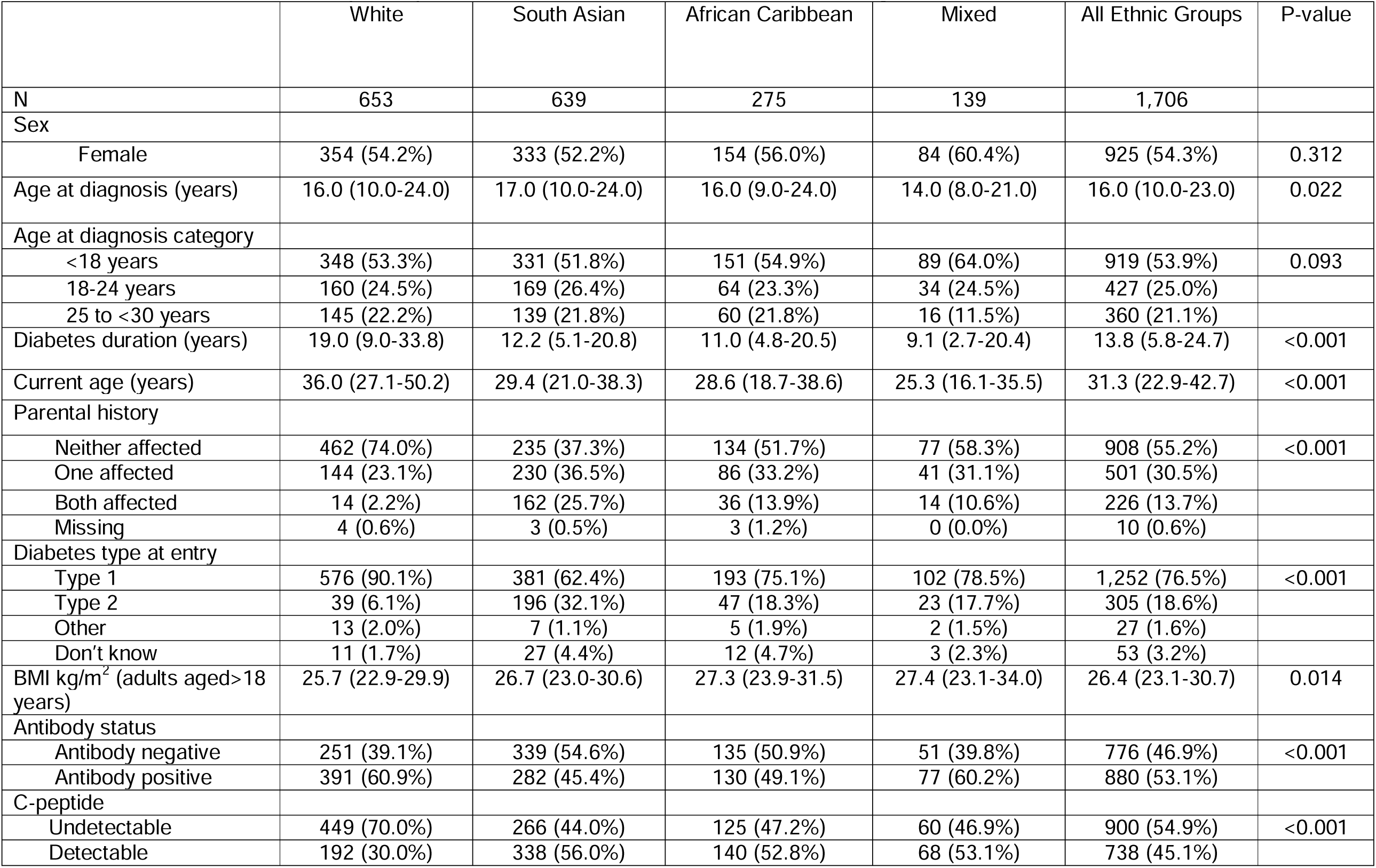
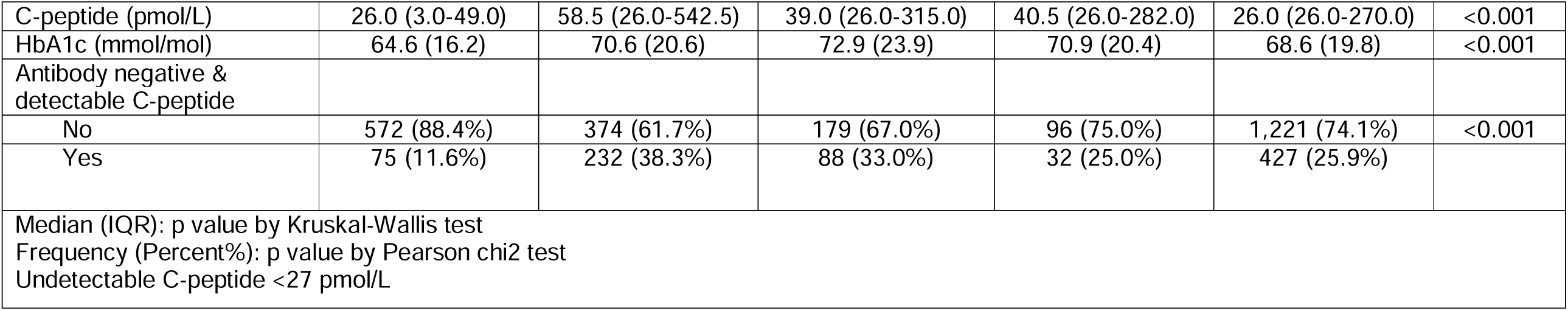
Characteristics of all participants recruited to the MY DIABETES study.

### Biomarker stratification

Of 1,706 participants, 900 had an undetectable C-peptide and of the 738 with detectable C-peptide, 303 were antibody positive. Therefore 427 individuals met study criteria for genetic testing (figure 1).

These criteria identified significantly different proportions across ethnic groups (table 1); C-peptide levels were undetectable in 449 (70.0%) of White participants, 266 (44.0%) of South Asian, 125 (47.2%) of African Caribbean and 60 (46.9%) of those from Mixed ethnic groups (p<0.001). In those with detectable C-peptide (>27 pmol/L), antibody positivity was encountered in 117 (61%) of White, 106 (31.0%) South Asian, 52 (38.0%) of African-Caribbean and 36 (53%) of mixed ethnicity p<0.001). Proportions meeting broad study criteria for genetic testing (antibody negative with detectable C-peptide) were also significantly different by ethnicity with 11.6 % (75/653) of White, 36.0% (232/639) South Asian, 31.0% (86/275) of African-Caribbean and 23% (32/139) of Mixed ethnicity p<0.001.

### Cohort prevalence & Detection rate

In total 39 cases of monogenic diabetes were identified in probands of which 25 were new diagnoses and 14 were known. The duration of diabetes in those newly diagnosed with monogenic diabetes was 9.1 (7.2-30.5) in White, 12.4 (3.4-21.5) in South Asian, 11.2 (1.5-32.7) in African-Caribbean and 9.1 (3.4-21.6) years in those from mixed ethnicity groups (p=0.75).

As a proportion of all participants recruited (minimum cohort prevalence), monogenic diabetes was found in 2.3% (39/1,706) of all participants and did not vary significantly by ethnicity (figure 1); 2.1% in White, 2.0% in South Asian, 2.5% in African-Caribbean and 3.6% in Mixed ethnic groups (p=0.70).

Applying these broadest criteria for genetic testing (antibody negative with detectable C-peptide), the detection rate for monogenic diabetes varied significantly by ethnic group; 17.7% (14/79) in White, 5.3% (13/243) in South Asian, 8.0% (7/91) in African-Caribbean and 15.2% (5/33) in those with mixed ethnicity p<0.001 (table 2). The proportion of new cases of monogenic diabetes (as a fraction of all the cases) identified in each ethnic group was; 43% (6/14) in White, 77% (10/13) in South Asian, 57% (4/7) in African-Caribbean and 100% (5/5) in mixed ethnicities, p=0.09. Compared to White participants, a new diagnosis of MODY (as a proportion of all MODY cases) was higher in non-White participants (79% vs 44%, p=0.019). The number needed to test to identify one monogenic case in each ethnic group using antibody negativity and a detectable C-peptide as the sole criteria, was 6, 19, 13 and 7 in White, South Asian, African-Caribbean and Mixed ethnic groups, respectively.

**Table 2:** Characteristics of those with and without pathogenic MODY mutations within each ethnic group studied.

|  | White |  |  | South Asian |  |  | African or Caribbean |  |  | Mixed |  |  |
| --- | --- | --- | --- | --- | --- | --- | --- | --- | --- | --- | --- | --- |
|  | No mutation | Mutation | P-value | No mutation | Mutation | P-value | No mutation | Mutation | P-value | No mutation | Mutation | P-value |
| N | 59 | 14 |  | 211 | 13 |  | 76 | 9 |  | 27 | 5 |  |
| **Sex** |  |  |  |  |  |  |  |  |  |  |  |  |
| Female | 38<br>(64.41%) | 7<br>(50.00%) | 0.319 | 111<br>(52.61%) | 10 (76.92%) | 0.088 | 46 (60.53%) | 6 (66.67%) | 0.721 | 18<br>(66.67%) | 4 (80.00%) | 0.555 |
| Male | 21<br>(35.59%) | 7<br>(50.00%) |  | 100<br>(47.39%) | 3 (23.08%) |  | 30 (39.47%) | 3 (33.33%) |  | 9<br>(33.33%) | 1 (20.00%) |  |
| **Current Age (years)** | 34.79<br>(28.62-45.53) | 31.91<br>(29.04-49.52) | 0.769 | 32.41<br>(24.77-41.27) | 23.12<br>(18.59-35.63) | 0.171 | 30.38<br>(22.51-42.38) | 28.99<br>(23.90-37.89) | 0.989 | 29.11<br>(23.09-43.04) | 20.90 (15.67-32.41) | 0.421 |
| **Age diagnosis category** |  |  |  |  |  |  |  |  |  |  |  |  |
| <18 years | 9<br>(15.25%) | 5<br>(35.71%) | 0.017 | 62<br>(29.38%) | 8 (61.54%) | 0.049 | 27 (35.53%) | 2 (22.22%) | 0.031 | 12<br>(44.44%) | 3 (60.00%) | 0.436 |
| 18-24 years | 22<br>(37.29%) | 8<br>(57.14%) |  | 77<br>(36.49%) | 2 (15.38%) |  | 19 (25.00%) | 6 (66.67%) |  | 8<br>(29.63%) | 2 (40.00%) |  |
| 25 to <30 years | 28<br>(47.46%) | 1 (7.14%) |  | 72<br>(34.12%) | 3 (23.08%) |  | 30 (39.47%) | 1 (11.11%) |  | 7<br>(25.93%) | 0 (0.00%) |  |
| **Age at diagnosis (years)** | 23.00<br>(19.00-27.00) | 18.00<br>(14.00-19.00) | 0.001 | 22.00<br>(16.00-26.00) | 15.00<br>(13.00-20.00) | 0.012 | 20.00<br>(14.40-26.00) | 21.00<br>(18.00-23.00) | 0.875 | 18.00<br>(14.00-25.00) | 15.00 (12.00-18.00) | 0.139 |
| **Diabetes duration (years)** | 12.69<br>(3.57-21.93) | 20.31<br>(8.71-30.52) | 0.062 | 11.28<br>(4.09-18.34) | 10.63 (3.39-20.48) | 0.988 | 9.77 (2.46-18.78) | 7.73 (5.51-16.89) | 0.842 | 10.12<br>(2.15-20.04) | 8.90 (1.46-13.41) | 0.697 |
| **Parental history of diabetes** |  |  |  |  |  |  |  |  |  |  |  |  |
| Neither parent | 24<br>(42.86%) | 3<br>(21.43%) | 0.343 | 34<br>(16.19%) | 5 (38.46%) | 0.042 | 28 (37.84%) | 1 (12.50%) | 0.309 | 6<br>(23.08%) | 2 (50.00%) | 0.512 |
| One parent | 25<br>(44.64%) | 10<br>(71.43%) |  | 79<br>(37.62%) | 6 (46.15%) |  | 30 (40.54%) | 6 (75.00%) |  | 12<br>(46.15%) | 1 (25.00%) |  |
| Both parents | 6<br>(10.71%) | 1 (7.14%) |  | 97<br>(46.19%) | 2 (15.38%) |  | 15 (20.27%) | 1 (12.50%) |  | 8<br>(30.77%) | 1 (25.00%) |  |
| **Initial Treatment** |  |  |  |  |  |  |  |  |  |  |  |  |
| Diet only | 14<br>(23.73%) | 3<br>(23.08%) | 0.815 | 30<br>(14.42%) | 1 (7.69%) | 0.305 | 7 (9.33%) | 3 (33.33%) | 0.155 | 0 (0.00%) | 2 (40.00%) | 0.007 |
| Tablets | 23<br>(38.98%) | 4<br>(30.77%) |  | 94<br>(45.19%) | 5 (38.46%) |  | 23 (30.67%) | 3 (33.33%) |  | 12<br>(44.44%) | 1 (20.00%) |  |
| Insulin | 22<br>(37.29%) | 6<br>(46.15%) |  | 65<br>(31.25%) | 7 (53.85%) |  | 38 (50.67%) | 3 (33.33%) |  | 12<br>(44.44%) | 2 (40.00%) |  |
| Insulin & tablets |  |  |  | 19<br>(9.13%) | 0 (0.00%) |  | 7 (9.33%) | 0 (0.00%) |  | 3<br>(11.11%) | 0 (0.00%) |  |
| **Currently on insulin?** |  |  |  |  |  |  |  |  |  |  |  |  |
| Yes | 39<br>(66.10%) | 4<br>(28.57%) |  | 145<br>(68.72%) | 8 (61.54%) |  | 58 (76.32%) | 4 (44.44%) |  | 21<br>(77.78%) | 4 (80.00%) | 0.912 |
| **BMI kg/m2 (age >18 yrs)** | 31.57<br>(26.16-37.46) | 23.59<br>(21.86-25.80) | <0.001 | 28.64<br>(24.68-33.24) | 22.23<br>(20.98-27.25) | 0.001 | 29.41<br>(24.78-33.52) | 26.19<br>(23.67-34.89) | 0.944 | 33.99<br>(27.91-40.65) | 31.56 (21.68-34.88) | 0.356 |
| **Waist:Hip Ratio** | 0.93<br>(0.85-0.96) | 0.87<br>(0.84-0.94) | 0.170 | 0.92<br>(0.88-0.97) | 0.86 (0.80-0.97) | 0.105 | 0.91 (0.86-0.95) | 0.86 (0.83-0.96) | 0.218 | 0.93<br>(0.87-0.97) | 0.78 (0.73-0.89) | 0.061 |
| **C-peptide (pmol/L)** | 577.00<br>(156.00-864.00) | 339.00<br>(232.00-660.00) | 0.278 | 612.00<br>(301.00-1039.00) | 480.00<br>(386.00-712.00) | 0.276 | 386.50<br>(203.00-660.50) | 390.00<br>(362.00-501.00) | 0.825 | 349.00<br>(228.00-695.00) | 222.00 (214.00-585.00) | 0.421 |
| **HbA1c (mmol/mol)** | 58.00<br>(46.00-77.00) | 54.50<br>(48.00-62.50) | 0.251 | 67.00<br>(53.00-84.00) | 57.00<br>(48.50-71.00) | 0.164 | 70.00<br>(52.00-91.00) | 63.50<br>(50.50-83.50) | 0.736 | 71.00<br>(46.00-83.00) | 55.00 (50.00-64.00) | 0.191 |
| **HDL (mmol/L)** | 1.20<br>(0.97-1.40) | 1.51<br>(1.24-1.90) | 0.003 | 1.10<br>(0.93-1.30) | 1.25 (1.11-1.54) | 0.061 | 1.34 (1.06-1.67) | 1.70 (1.19-1.80) | 0.312 | 1.23<br>(1.00-1.63) | 1.58 (1.42-1.60) | 0.384 |
| **Triglycerides (mmol/L)** | 1.40<br>(0.91-1.93) | 1.45<br>(0.74-1.92) | 0.763 | 1.52<br>(1.14-2.37) | 1.02 (0.90-1.60) | 0.043 | 0.93 (0.60-1.43) | 0.89 (0.70-0.96) | 0.777 | 1.37<br>(0.79-1.89) | 0.82 (0.62-1.14) | 0.149 |

Median (IQR): p value by Kruskal-Wallis test
Frequency (Percent%): p value by Pearson chi2 test
In each ethnicity the 'No mutation' group comprises people who were tested for monogenic diabetes but no mutation was found, excludes variants of unknown significance

### MODY phenotype

MODY phenotype was similar across ethnic groups (supplementary table 1) with no significant difference in the typical features of monogenic diabetes including age at diagnosis, parental history of diabetes, proportion initially non-insulin treated. Median BMI was also similar irrespective of ethnicity though a tendency towards higher BMI was observed in African-Caribbean and mixed ethnic groups. The proportion of monogenic diabetes cases with BMI >30 kg/m^2^ out of all cases found was 7% (1/14) in White, 8% (1/13) in South Asian, 14% (1/7) in African-Caribbean and 40% (2/5) in Mixed ethnicity groups.

In White and African-Caribbean participants with monogenic diabetes, over 70% had a one parent affected with diabetes, but in South Asian participants, 46% had one parent affected and 15% had both parents affected.

Affected genes ranged from common MODY genes (HNF1A, HNF4A, GCK and HNF1B), mitochondrial mutations to rarer genes (supplementary table 2). 100% of White, 62% of South Asian, 71% of African-Caribbean and 80% of Mixed ethnicity participants harboured mutations in HNF1A, HNF4A, GCK and HNF1B genes or mitochondrial mutations.

### Age-at-diagnosis, parental history & BMI differences

Analysing individuals who were tested for monogenic diabetes, but found not to harbour mutations (mutation negative), across ethnic groups, BMI was significant lower in the South Asian group 28.6 kg/m^2^ (24.8-32.6) vs White 31.6 (26.1-37.3), African-Caribbean 29.5 (24.8 – 33.9) and Mixed 33.4 kg/m^2^ (28-40.5), p=0.003. The South Asian participants were most likely to have both parents affected with diabetes: 45.8% of South Asian participants who tested mutation negative had both parents affected versus 10% in White, 20% in Black and 30% in mixed participants, p<0.001 (Supplementary table 3 & supplementary figure 1).

Within White ethnicity participants, those without mutations in diabetes genes had a significantly higher age at diagnosis than those with mutations (25 vs 18 years, p<0.001) (table 2). Similarly South Asian participants without mutations were significantly older than those with (23 vs 17 years, p=0.02), however no age difference was observed in African-Caribbean or Mixed ethnicity participants with and without MODY (figure 2).

**Figure 2:**
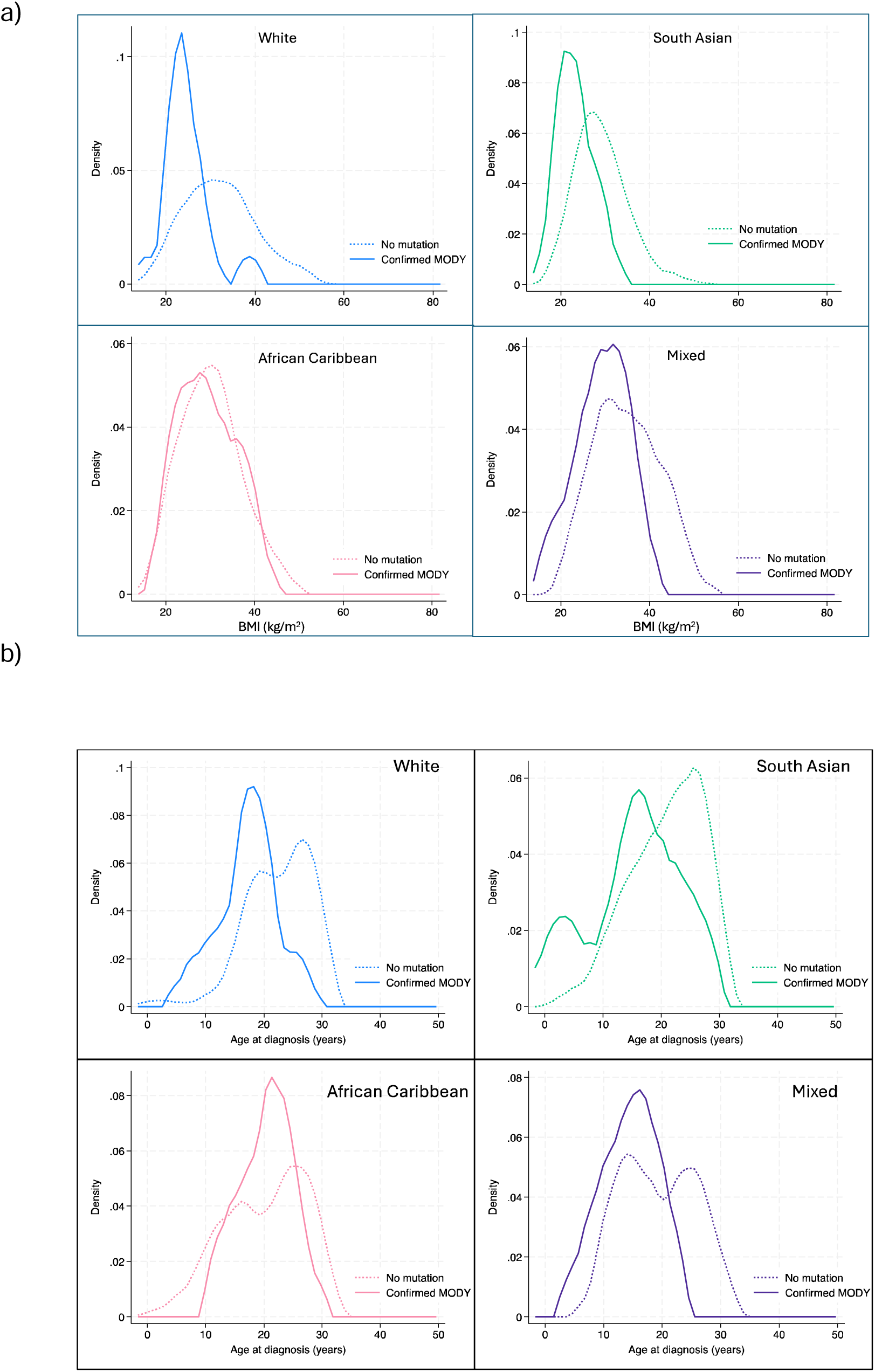
BMI (a) and age-at -diagnosis (b) distribution of those with confirmed pathogenic mutations versus those in whom genetic testing was undertaken but no mutation was found, by ethnicity

For South Asian participants without monogenic diabetes who had been tested, the proportions with both parents affected rose to 46%. However, in over a third of cases no parental history was present, which was partially explained by recessive monogenic diabetes mutations found.

BMI was significantly lower in White participants who had MODY vs those who didn’t (23.6 vs 31.6 kg/m^2^ p<0.001) and in South Asian participants (22.3 vs 28.6 p=0.001). In African-Caribbean and Mixed ethnicity participants, BMI was non-significantly different in those with MODY vs without (26.2 vs 29.4 kg/m^2^ p=0.94 and 34.0 vs 31.6 p=0.36, respectively) (figure 2).

The distribution of BMI across ethnic groups in those tested for monogenic diabetes, varied significantly by ethnic group (figure 3). A greater proportion of South Asian and African-Caribbean individuals who were found not to have monogenic diabetes were in a normal BMI weight category; 17% of White, 28% of South Asian and 32% of African-Caribbean participants had a BMI <25 kg/m^2^. In each ethnic group the majority of people tested had a BMI >30 kg/m^2^. However, notably in each ethnic group, monogenic diabetes cases were also found in higher BMI categories.

**Figure 3:**
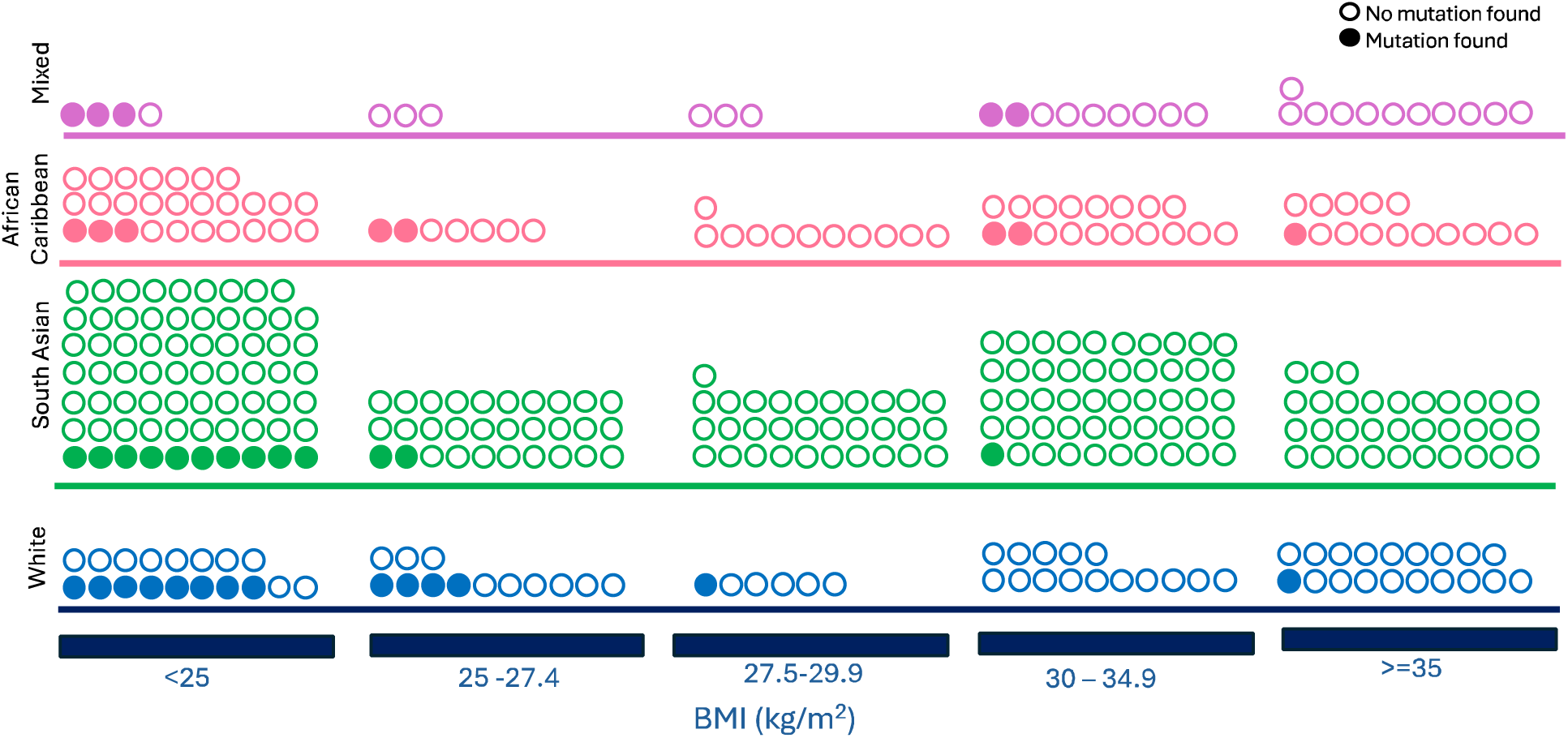
Schematic visualisation of BMI in individual participants with and without pathogenic mutations, by ethnicity. This plot shows the individual genetic testing results in participants who had detectable C-peptide and were pancreatic antibody negative. Filled circles indicate that a pathogenic mutation was found and unfilled means no mutation was found. Different colours indicate different ethnic groups, as labelled.

### Effectiveness of BMI criteria

Applying variable cut-offs for BMI led to differences in detection rate that varied across ethnic group (table 3). Specific cut-offs were selected based on the distribution of cases by BMI category in each ethnic group (figure 2a). In White individuals a cut-off that included participants with a BMI <30 kg/m^2^ achieved a detection rate of 35%, missing a single case (false negative rate 7%) and reduced the number needed to test from 6 (without any BMI criteria) to 3.

**Table 3:**
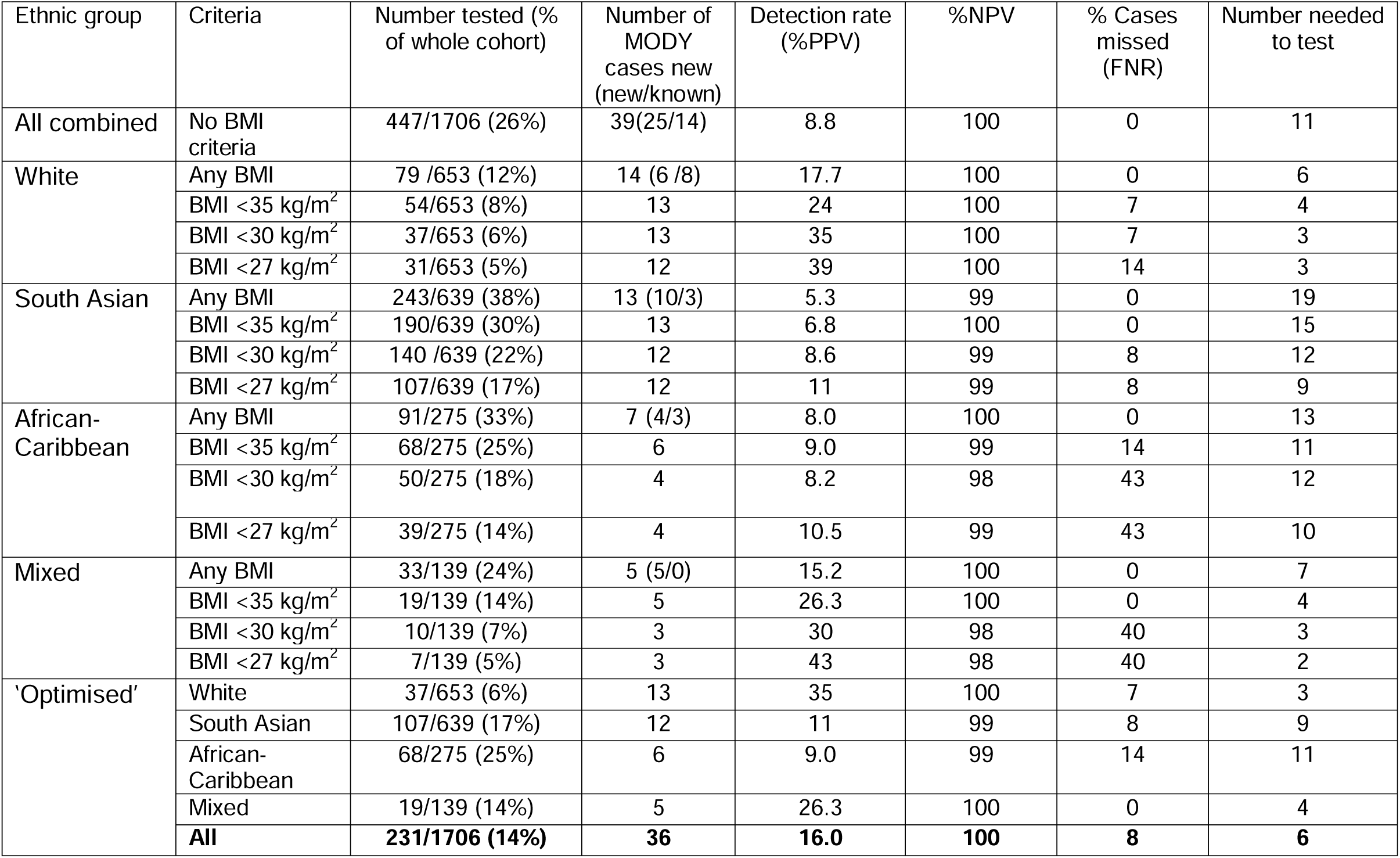
Effect of different BMI criteria on detection of pathogenic MODY mutations in each ethnic group in those individuals with detectable C-peptide with pancreatic antibodies negative.

The same BMI cut-off in South Asian people however achieved only an 8.6% detection rate with number needed to test of 12 patients. By reducing the BMI cut-off to <27.5 kg/m^2^ the detection rate increased to 11% and the number needed to test reduced to 9 patients.

A lower BMI cut off of <27.5 kg/m^2^ in African-Caribbean and Mixed ethnicity participants achieved the highest detection rate (10.5% and 43%), but missed 43 and 40% of cases, respectively. A higher BMI cut off <35 kg/m^2^ achieved a detection rate of 9% in African-Caribbean and 26.3% in Mixed ethnicity participants, with a false negative rate of 14% in African-Caribbean and 0% in Mixed and with 11 and 4 patients needed to test, respectively. By applying ‘optimised’ BMI cut-off for each individual ethnicity (<30 kg/m^2^ in White, <27.5 in South Asian and <35 in African-Caribbean and Mixed groups), the overall cohort detection rate increased from 8.8% to 16% with a false negative rate of 8% (3/39 cases missed) and with a number needed to test of 6 patients to identify 1 monogenic diabetes case.

External validation

Ethnicity specific BMI cut-offs applied to the national genetics referral registry demonstrated a similar trend in detection rates across ethnic groups (supplementary figure 2). The overall detection rate for monogenic diabetes was 30% in White (2,713 mutations found / 9019 cases tested), 11% in South Asian (144/1257) and 10% in Black (36/449). By applying a BMI cut off of 27 kg/m^2^ in the South Asian ethnic group, the detection rate increased to 14%. The number needed to test in South Asian was 8.7 without a BMI threshold and reduced to 6.9 at a cut-off of 27 kg/m^2^ (figure 4). However, the detection rate at a cut-off of 30 kg/m^2^ in South Asian was not dissimilar at 13%. As the registry is biased towards leaner BMI individuals that have been referred, we used population data to estimate the increase in referrals received if the BMI threshold was <27 kg/m^2^ compared to <30 kg/m^2^.

**Figure 4:**
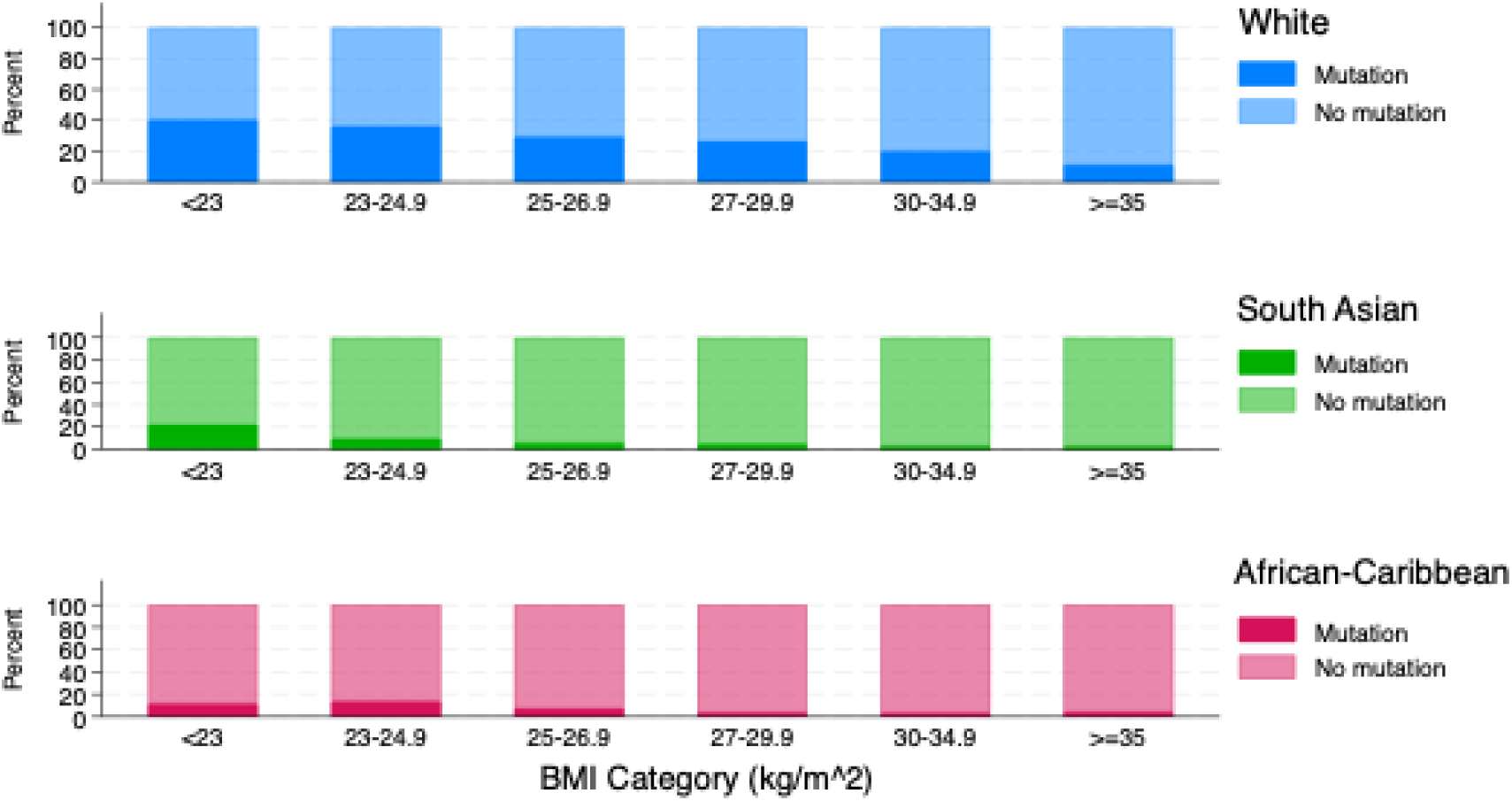
Proportion of pathogenic mutations found out of those tested in the National Genetic Laboratory Registry (Exeter) in each BMI category stratified by ethnicity

Local population data (supplementary table 4) showed that a cut-off of <30 kg/m^2^ would lead to 943 South Asian people diagnosed with type 2 diabetes under 30 years, being referred for testing – in total 51.7% of all South Asian individuals with type 2 diabetes diagnosed below 30 years (943/1823). At a cut off of 27kgm/^2^ this number reduced to 584 representing 32% of those with early-onset type 2 diabetes and reducing the numbers tested by 38%. In contrast, for White and African-Caribbean ethnic groups, the extra number of individuals referred between a BMI of 27 and 30 kg/m^2^ were 110 (12.7%) of all individuals with type 2 diabetes diagnosed <30 years and 73 (12%), respectively.

## Discussion

In line with a precision medicine approach, our analysis demonstrates the need for ethnic specific criteria to optimise monogenic diabetes detection. In participants with diabetes diagnosed <30 years recruited from hospital-based diabetes services, although a similar minimum cohort prevalence of monogenic diabetes was observed across all four ethnic groups, most monogenic diabetes cases were newly identified in South Asian, African-Caribbean and Mixed ethnicity participants. These newly identified cases were detected, on average, ∼10 years after the initial diagnosis of diabetes, demonstrating that current approaches work poorly in these under-represented ethnic groups.

Applying broad biomarker criteria to select for genetic testing worked well in White participants, minimising the number needed to test. In non-White groups, these criteria identified many misdiagnosed MODY cases, eliminating clinical bias that has previously precluded genetic testing, but also selected more participants for testing due to the higher prevalence of early-onset type 2 diabetes in these populations. This variable prevalence lowered the detection rate as the criteria did not differentiate between type 2 diabetes and MODY, leading to over-testing.

We showed that to optimise detection of monogenic diabetes, tailored BMI cut-offs applied to each ethnic group (differentiating early-onset type 2 diabetes which is associated with obesity from MODY) can increase the detection rate and reduce the numbers needed to test to identify one case. By individualising criteria for participants based on their ethnicity, the overall cohort detection rate doubled from 8% to 16%, requiring just 6 people in a multiethnic population to be tested, to identify 1 case (from 11 using broad criteria). However, applying the same criteria to all ethnic groups led to under-testing and missed cases.

Our external validation, provides additional data to support a cut-off of 27 kg/m^2^ in South Asian and 30 kg/m^2^ in African-Caribbean participants, based on optimal detection rates offset against false negative rates and with population data showing the impact on referral nunbers.

In summary, applying biomarker criteria to stratify participants for genetic testing can eliminate clinical bias in selection for genetic testing across diverse ancestry groups and improve diagnosis of monogenic diabetes in all ethnic groups studied and the detection rate can be optimised by the application of tailored ethnic-specific BMI cut offs for each ancestry group. Therefore, a precision-based approach to the diagnosis of monogenic diabetes can reduce error in diagnoses, become more cost-effective my reducing numbers needed to test and highlights the need to study diverse ancestries when developing diagnostic pathways.

To our knowledge this is the first study to systematically apply biomarker criteria for the detection of monogenic diabetes in unselected patients from three different ethnic groups and to determine optimised criteria for monogenic diabetes detection, tailored to specific ethnic groups.

Several studies have used biomarker-based approaches to diagnose monogenic diabetes. The United study ^1^ reported a 3.6% prevalence in a predominantly White cohort, slightly higher than our study’s 2.1% to 3.6% across ancestry groups, suggesting a potentially higher population prevalence. Unlike the United study, which only tested individuals with C-peptide >200 pmol/L, our protocol included any detectable C-peptide (>27 pmol/L) to avoid bias in non-White populations. Five monogenic diabetes cases in our study had C-peptide <200 pmol/L and would have been missed with higher thresholds, highlighting variability in C-peptide levels across ethnicities.

The PRODIGY ^13^ study found a 2.8% detection rate in youth with overweight or obesity labelled as type 2 diabetes, but excluded lean participants and did not report ethnicity. This is lower than our 8.8% detection rate across ethnicities without BMI criteria, similar to the SEARCH study ^9^, which found an 8.0% detection rate by applying biomarker criteria regardless of body weight.

An Oxford study of mostly White patients with type 2 diabetes ^14^ diagnosed before age 30 reported a 25% detection rate for monogenic diabetes, testing only HNF1A/HNF4A genes. This is higher than our 17.7% rate in White participants without BMI criteria, likely due to our inclusion of any detectable C-peptide and diagnosis-agnostic criteria, potentially incorporating patients with type 1 diabetes still producing low C-peptide.

Few studies report monogenic diabetes detection rates across diverse ancestries. A French study^8^ using sequential data from a central genetic testing laboratory found variable detection rates. Unlike our study, their cohort comprised of pre-selected patients in whom referring clinicians already suspected monogenic diabetes, upon which they applied further criteria such as family history, age-at-diagnosis, and BMI cut-offs to select for genetic testing. In this highly selected cohort, the detection rate was 23.6% in White European, 11.8% in African and 7.3% in South Asian patients. They found that applying BMI thresholds (<26 kg/m² in Caribbean, <29 kg/m² in African, and <23.9 kg/m² in South Asian patients) improved detection rates. The higher detection rates and lower BMI cut-offs in this study likely reflect the highly pre-selected population referred for testing. The present analysis, conducted in an unselected population recruited from clinics without suspected MODY, is more representative of typical patient encounters, demonstrates that pathogenic mutations are observed across a broader spectrum of BMI values.

A previous analysis of the centralised UK genetic testing laboratory database showed a detection rate of 12.5% in UK South Asian people, significantly lower than the White detection rate (29%) ^7^ but higher than that observed in the present study. Again, reflecting that the present study was conducted in an unselected population.

Traditional factors such as age-at-diagnosis and family history showed variable utility in separating early-onset type 2 diabetes from monogenic diabetes in this analysis. South Asian participants with monogenic diabetes had a significantly lower age-at-diagnosis (than those without), but this may have been ‘lowered’ due to the range of affected genes found in this cohort, which included a new diagnosis of Wolframs syndrome and two rare recessive cases in ZNF808 and SLC19A2 presenting in children. A higher rate of recessive mutations has been observed in populations with greater consanguinity^15^.

Over 80% of South Asian participants without monogenic diabetes had one or both parents affected with diabetes (vs ∼ 60% in those with monogenic diabetes), making a parental history of diabetes a less sensitive discriminator. However, a history of both parents affected, occurred more frequently in people without monogenic diabetes (46% vs 16%), suggesting that using one vs both parents could have discriminatory value as both parents affected maybe more common in early-onset type 2 diabetes^16^.

Our study confirms that phenotype of monogenic diabetes is broadly similar across 4 ethnic groups. This confirms previous findings from other studies^7,8^ and indicates that the higher proportion of misclassified monogenic diabetes diagnoses in non-White participants, is not down to an atypical monogenic diabetes phenotype, rather, the difference reflects variation in the phenotype of participants found to *not* harbour monogenic diabetes. We show that whilst clinical features vary considerably between White participants (undergoing genetic testing) with and without monogenic diabetes e.g. BMI 24 vs 32 kg/m, these differences are attenuated in South Asian and absent in African-Caribbean and mixed ethnicity participants signifying that the phenotype of early-onset type 2 diabetes is less distinguishable from monogenic diabetes in non-White ethnic groups. Early-onset type 2 diabetes is strongly associated with obesity in epidemiological studies^17,18^, contrasting studies of older-onset type 2 diabetes in which South Asian patients had much lower BMI than White^19^. In our analysis ∼30% of south Asian and African-Caribbean participants had a BMI <25 kg/m^2^, but did not harbour a monogenic diabetes mutation. The diagnosis in this group is unclear and could be autoimmune type 1 diabetes with absent pancreatic autoantibodies, monogenic diabetes in an unknown gene or indeed lean type 2 diabetes. South Asian patients with early-onset type 2 diabetes in India were found to have a mean BMI of ∼27 kg/m^2^ in both selected clinic-based studies^20^ and population-level analysis^21^.

The BMI cut-offs identified in the present analysis are supported by analyses proposing revised ethnicity-specific cut-offs for BMI that define obesity across the population^22^ showing a higher risk of type 2 diabetes with BMI >23·9 kg/m^2^ and >28.1 kg/m^2^ in Asian and Black populations, respectively in the UK.

In the present study monogenic diabetes was found in all BMI categories, however, the number needed to test to identify one case and associated cost of testing in a BMI category with low probability, makes universal testing across BMI groups unfeasible.

It is notable that in the present study, higher BMI monogenic diabetes was predominantly observed in African-Caribbean and Mixed ethnicity cases. Although the number of MODY cases is small and requires confirmation, this finding may reflect well-documented differences in body composition^23^, including a higher proportion of lean muscle mass and different fat distribution patterns in African-Caribbean populations^24^. Additionally, African-Caribbean populations exhibit higher rates of obesity, which has been consistently reported in epidemiological studies^25,26^.

Our findings need confirmation in other unselected cohorts of similar and diverse ethnic backgrounds. However, the lack of multiethnic or non-White European cohorts highlights the need for future research to diversify cohorts and report subethnic groups to enable combined analysis. The MODY probability calculator ^27^, developed in a White population, offers another method for improving monogenic diabetes detection, and future validation in other ethnic groups is essential. Importantly, the goal of ethnic-specific criteria should not be to match White detection rates, but to maximise case identification while minimising unnecessary testing, given the varying prevalence of early-onset type 2 diabetes.

### Limitations

Our study has some limitations. First, the MY DIABETES study recruited participants from hospital clinics and is not a population level sample, therefore we are unable to conclusively report prevalence of monogenic diabetes across ethnic groups. We also only classified ethnicity using self-reported data. Our study comprises cross-sectional data and we do not have access to phenotype data at the point of diagnosis. It is possible that some of the variables analysed e.g. BMI may have changed over time from diagnosis. However, as most monogenic diabetes cases are not diagnosed at the time of first presentation of diabetes use of these cross-sectional data are likely to be more representative of the time point at which patients are considered for genetic testing. We did not genetically test participants who were antibody positive or with undetectable C-peptide nor did we undertake whole genome or exome sequencing in our pipeline. It is therefore possible that some of the participants labelled as not having monogenic diabetes, do harbour monogenic diabetes. Finally, the African-Caribbean group was smaller than the South Asian and White ethnic groups, as recruitment started later and was prematurely stopped due to the pandemic.

## Conclusion

This study underscores the importance of applying tailored, ethnicity-specific criteria to optimise the detection of monogenic diabetes in diverse populations. By using biomarker-criteria and adjusting BMI thresholds across ethnic groups, we were able to significantly enhance detection rates and reduce numbers needed to test. We demonstrate that a precision medicine approach, eliminates clinical bias in selection for genetic testing and modifies diagnostic criteria to ancestral differences, is essential for improving the accuracy of monogenic diabetes diagnoses. Ultimately, a tailored approach is likely to improve cost-effectiveness of testing pathways. Our study, the first to systematically apply biomarker criteria across multiple ethnicities, highlights the need for greater inclusion of diverse ancestries in the development of diagnostic pathways to minimise misclassification and enhance patient care, equitably.

## Data Availability

In order to reduce the risk of re-identification, de-identified patient-level clinical data are available under controlled access. Researchers interested in obtaining anonymised individual participant data and study documents should submit requests to the corresponding author. These requests will be reviewed by the MY DIABETES study Steering Committee. Data access will be granted to academic teams with scientifically valid research questions and appropriate expertise. Approved data for release, will be shared under a relevant institutional data transfer agreement.

## Acknowledgements

We acknowledge the research participants living with diabetes who consented to participate in this study along with the local principal investigators and research team members at the 41 sites recruiting to the MY DIABETES study, listed in Supplementary material.

