## Supplementary material for "Precision diagnosis for monogenic diabetes requires ethnicity specific criteria for genetic testing"

**Supplementary table 1:** Characteristics of participants with all pathogenic MODY mutations found, across ethnic groups.

|  | **White** | **South Asian** | **African-Caribbean** | **Mixed** | **P-value** |
| --- | --- | --- | --- | --- | --- |
| **N** | 14 (34.1%) | 13 (31.7%) | 9 (22.0%) | 5 (12.2%) |  |
| **Sex** |  |  |  |  |  |
| Female | 7 (50.0%) | 10 (76.9%) | 6 (66.7%) | 4 (80.0%) | 0.437 |
| Male | 7 (50.0%) | 3 (23.1%) | 3 (33.3%) | 1 (20.0%) |  |
| **Current Age** | 31.9 (29.0-49.5) | 23.1 (18.6-35.6) | 29.0 (23.9-37.9) | 20.9 (15.7-32.4) | 0.266 |
| **Age at diagnosis (years)** | 18.0 (14.0-19.0) | 15.0 (13.0-20.0) | 21.0 (18.0-23.0) | 15.0 (12.0-18.0) | 0.206 |
| **Age at diagnosis category (%)** |  |  |  |  |  |
| <18 years | 5 (35.7%) | 8 (61.5%) | 2 (22.2%) | 3 (60.0%) | 0.196 |
| 18 to <25 years | 8 (57.1%) | 2 (15.4%) | 6 (66.7%) | 2 (40.0%) |  |
| 25 to <30 years | 1 (7.1%) | 3 (23.1%) | 1 (11.1%) | 0 (0.0%) |  |
| **Diabetes duration (years)** | 20.3 (8.7-30.5) | 10.6 (3.4-20.5) | 7.7 (5.5-16.9) | 8.9 (1.5-13.4) | 0.214 |
| **Known or new diagnosis** |  |  |  |  |  |
| New diagnosis | 6 (42.9%) | 10 (76.9%) | 6 (66.7%) | 5 (100.0%) | 0.086 |
| Known diagnosis | 8 (57.1%) | 3 (23.1%) | 3 (33.3%) | 0 (0.0%) |  |
| **Parental history of diabetes** |  |  |  |  |  |
| Neither parent affected | 3 (21.4%) | 5 (38.5%) | 1 (12.5%) | 2 (50.0%) | 0.566 |
| One parent affected | 10 (71.4%) | 6 (46.2%) | 6 (75.0%) | 1 (25.0%) |  |
| Both parents affected | 1 (7.1%) | 2 (15.4%) | 1 (12.5%) | 1 (25.0%) |  |
| **Initial Treatment** |  |  |  |  |  |
| Diet only | 3 (23.1%) | 1 (7.7%) | 3 (33.3%) | 2 (40.0%) | 0.764 |
| Tablets | 4 (30.8%) | 5 (38.5%) | 3 (33.3%) | 1 (20.0%) |  |
| Insulin | 6 (46.2%) | 7 (53.8%) | 3 (33.3%) | 2 (40.0%) |  |
| **Currently on insulin treatment** |  |  |  |  |  |
| No | 10 (71.4%) | 5 (38.5%) | 5 (55.6%) | 1 (20.0%) | 0.161 |
| Yes | 4 (28.6%) | 8 (61.5%) | 4 (44.4%) | 4 (80.0%) |  |
| **Current BMI kg/m^2^ (age >18 yrs)** | 23.6 (21.9-25.8) | 22.2 (21.0-27.3) | 26.2 (23.7-34.9) | 31.6 (21.7-34.9) | 0.087 |
| **Waist:Hip Ratio** | 0.9 (0.8-0.9) | 0.9 (0.8-1.0) | 0.9 (0.8-1.0) | 0.8 (0.7-0.9) | 0.505 |
| **C-peptide (pmol/L)** | 339.0 (232.0-660.0) | 480.0 (386.0-712.0) | 390.0 (362.0-501.0) | 222.0 (214.0-585.0) | 0.595 |
| **HbA1c (mmol/mol)** | 54.5 (48.0-62.5) | 57.0 (48.5-71.0) | 63.5 (50.5-83.5) | 55.0 (50.0-64.0) | 0.660 |
| **HDL (mmol/L)** | 1.5 (1.2-1.9) | 1.3 (1.1-1.5) | 1.7 (1.2-1.8) | 1.6 (1.4-1.6) | 0.221 |
| **Triglycerides (mmol/L)** | 1.4 (0.7-1.9) | 1.0 (0.9-1.6) | 0.9 (0.7-1.0) | 0.8 (0.6-1.1) | 0.303 |
| Data shown are n (%) or median (IQR) Median (IQR): p value by Kruskal-Wallis test Frequency (%): p value by Pearson chi2 test | | | | | |

**Supplementary Table 2:** All pathogenic proband mutations in monogenic diabetes genes identified as part of the study.

Het, heterozygous; homo, homozygous; maternal = mat.

*Due to the rarity of some mutations, clinical information has been de-identifed

| Ethnicity | Gene | Amino acid change | Diagnosis status | Inheritance | Age at study (yrs) | Age at diagnosis (yrs) | Duration (yrs) | Currently insulin-treated | Parent affected | BMI (kg/m^2^) | WHR | C-peptide (pmol/L) | HbA1c (mmol/mol) |
| --- | --- | --- | --- | --- | --- | --- | --- | --- | --- | --- | --- | --- | --- |
| African or Caribbean | GCK | p.Asp217Glu | New | Het | 17 | 14 | 3 | No | Both | 21 | 0.79 | 389 | 44 |
| African or Caribbean | ABCC8 | p.Thr1516Met | New | Het | 22 | 14 | 8 | Yes | One | 35 | 0.78 | 390 | 84 |
| African or Caribbean | PPARG | p.Phe415Leufs*15 | Known | Het | 46 | 20 | 26 | Yes |  | 26 | 0.96 | 501 |  |
| African or Caribbean | Mitochondrial | 3243 A>G 10% heteroplasmy | New | Mat | 33 | 27 | 6 | No | One | 34 | 0.96 | 572 | 83 |
| African or Caribbean | HNF1A |  | Known | Het | 38 | 21 | 17 | No | One |  | 0.90 | 461 | 53 |
| African or Caribbean | HNF1A |  | Known | Het | 29 | 23 | 6 | No | One | 23 | 0.83 | 283 | 50 |
| African or Caribbean | Mitochondrial | 3243A>G (36% heteroplasmy) | New | Mat | 24 | 22 | 2 | Yes | Neither | 26 | 0.83 | 593 | 51 |
| Mixed | HNF1A | p.Leu144Pro | New | Het | 32 | 19 | 13 | Yes |  | 32 | 0.78 | 48 | 64 |
| Mixed | HNF4A | p.Arg114Gln | New | Het | 60 | 18 | 42 | Yes | Both | 35 | 0.89 | 222 | 70 |
| Mixed | GCK | p.Arg397Leu | New | Het | 9 | 8 | 1 | Yes | Neither |  |  | 214 | 43 |
| Mixed | GATA6 | p.Trp225* | New | Het | 21 | 12 | 9 | Yes | Neither | 22 | 0.73 | 585 | 55 |
| Mixed | HNF1A | p.Arg263His | New | Het | 16 | 15 | 1 | No | One |  |  | 692 | 50 |
| South Asian | ABCC8 | p.Ala1537Val | New | Het | 70 | 25 | 45 | Yes | One | 23 | 0.99 | 319 | 70 |
| South Asian | WFS1 | p.Val434del | New | Homo | 19 | 6 | 13 | Yes | Neither | 27 | 0.94 | 30 | 79 |
| South Asian | HNF1A | NM_000545.6:c.1501_1G>A splicing mutation | New | Het | 35 | 15 | 20 | Yes | One | 27 | 0.90 | 1169 | 87 |
| South Asian | Mitochondrial | 3243A>G | New | Mat | 23 | 17 | 6 | Yes | One | 21 | 1.00 | 614 | 72 |
| South Asian | GCK | p.Arg43_Gly44del | Known | Het | 36 | 25 | 11 | No | Both | 25 | 0.84 | 516 | 40 |
| South Asian | HNF1B | Whole gene deletion | New | Het | 36 | 25 | 11 | No | Both | 20 | 0.71 | 409 | 61 |
| South Asian | ZNF808 | del ex4-5 | Known | Homo | <10 | <10 | ~ | Yes | ~ |  |  | 54 | 46 |
| South Asian | SLC19A2 | p.Glu66Ter(p.E66*) | Known | Homo | <10 | <10 | ~ | Yes | ~ |  |  | 386 |  |
| South Asian | HNF4A | p.Val180Ile | New | Het | 23 | 20 | 3 | No | One | 21 | 0.80 | 388 | 53 |
| South Asian | GCK | p.Arg397Leu | New | Het | 18 | 15 | 3 | Yes | Neither | 18 | 0.73 | 903 | 53 |
| South Asian | HNF4A | Del exon 1 to promoter (g.42983993_42984592 del) chr 20 | New | Het | 35 | 13 | 22 | Yes | One | 31 | 0.82 | 480 | 50 |
| South Asian | KCNJ11 | P.Glu229Lys | New | Het | 57 | 15 | 42 | No | Neither | 22 | 0.97 | 747 | 61 |
| South Asian | GCK | p.Met251Thr | New | Het | 19 | 18 | 1 | No | One | 21 | 0.86 | 712 | 47 |
| White | GCK | p.Leu164Pro | New | Het | 32 | 10 | 22 | No | One | 22 |  | 232 | 47 |
| White | Mitochondrial | 3243A>G | Known | Mat | 35 | 16 | 19 | Yes | One | 26 | 1.00 | 42 | 72 |
| White | HNF1B | whole gene deletion | New | Het | 29 | 26 | 3 | No | Neither | 16 | 0.89 | 167 | 49 |
| White | HNF1A | p.Pro112Leu | New | Het | 50 | 19 | 31 | No | One | 25 | 0.97 | 293 | 33 |
| White | HNF1A | p.Ala251Tyr | New | Het | 27 | 18 | 9 | Yes | One | 28 | 0.92 | 93 | 54 |
| White | HNF1A | p.Ala116Val | Known | Het | 48 | 20 | 28 | No | Both | 22 | 0.84 | 628 | 64 |
| White | HNF4A | p.Gln268Ter | Known | Het | 68 | 19 | 49 | No | One | 39 | 0.85 | 713 | 61 |
| White | HNF4A | p.Arg290His | Known | Het | 27 | 18 | 9 | Yes | One | 22 | 0.79 | 385 | 74 |
| White | HNF1A | c.42_51delinsTG/N | Known | Het | 23 | 18 | 5 | No | Neither | 25 | 0.94 | 754 |  |
| White | GCK | p.Thr149Ile | Known | Het | 30 | 13 | 17 | No | Neither | 21 | 0.82 | 280 | 39 |
| White | HNF1A | p.Arg229Pro | Known | Het | 32 | 7 | 25 | No | One | 27 | 0.87 | 756 | 56 |
| White | HNF1A | p.Arg263His | New | Het | 31 | 24 | 7 | No | One | 24 | 0.96 | 416 | 52 |
| White | HNF1A | p.Arg203His | New | Het | 69 | 19 | 50 | No | One | 23 | 0.79 | 660 |  |
| White | HNF1A | p.His143Tyr | Known | Het | 70 | 14 | 56 | Yes | One | 22 | 0.86 | 260 | 55 |

**Supplementary table 3:** Comparison of cases tested but found not to carry a pathogenic MODY mutation, after exclusion of those with undetectable C-peptide or one or more antibody positive.

|  | **White** | **South Asian** | **African or Caribbean** | **Mixed** | **P-value** |
| --- | --- | --- | --- | --- | --- |
| **N (% of total recruited)** | 61/663 (9.2%) | 215 /658 (32.7%) | 77 /284 (27.1%) | 28 /141 (19.9%) |  |
| **Sex** |  |  |  |  |  |
| Female | 40 (65.6%) | 113 (52.6%) | 47 (61.0%) | 19 (67.9%) | 0.145 |
| Male | 21 (34.4%) | 102 (47.4%) | 30 (39.0%) | 9 (32.1%) |  |
| **Current age (years)** | 34.8 (28.7-45.0) | 32.7 (24.8-41.3) | 30.7 (22.8-42.4) | 29.2 (24.0-42.0) | 0.085 |
| **Age at diagnosis (years)** | 24.0 (19.0-28.0) | 22.0 (16.8-26.0) | 20.0 (14.8-26.0) | 19.5 (14.0-25.5) | 0.067 |
| **Age at diagnosis category n (%)** |  |  |  |  |  |
| <18 years | 9 (14.8%) | 62 (28.8%) | 27 (35.1%) | 12 (42.9%) | 0.047 |
| 18 to <25 years | 22 (36.1%) | 77 (35.8%) | 19 (24.7%) | 8 (28.6%) |  |
| 25 to <30 years | 30 (49.2%) | 76 (35.3%) | 31 (40.3%) | 8 (28.6%) |  |
| **Diabetes duration (years)** | 12.7 (3.6-21.3) | 11.3 (4.1-18.3) | 10.3 (2.5-19.0) | 9.1 (2.1-19.0) | 0.889 |
| **Parental history of diabetes** |  |  |  |  |  |
| Neither parent affected | 24 (41.4%) | 34 (15.9%) | 28 (37.3%) | 7 (25.9%) | <0.001 |
| One parent affected | 27 (46.6%) | 81 (37.9%) | 31 (41.3%) | 12 (44.4%) |  |
| Both parents affected | 6 (10.3%) | 98 (45.8%) | 15 (20.0%) | 8 (29.6%) |  |
| Missing | 1 (1.7%) | 1 (0.5%) | 1 (1.3%) | 0 (0.0%) |  |
| **Initial Treatment** |  |  |  |  |  |
| Diet only | 14 (23.0%) | 32 (15.1%) | 7 (9.2%) | 0 (0.0%) | 0.007 |
| Tablets | 24 (39.3%) | 95 (44.8%) | 24 (31.6%) | 13 (46.4%) |  |
| Insulin | 23 (37.7%) | 66 (31.1%) | 38 (50.0%) | 12 (42.9%) |  |
| Tablets & Insulin | 0 (0.0%) | 19 (9.0%) | 7 (9.2%) | 3 (10.7%) |  |
| **Currently on insulin** |  |  |  |  |  |
| Yes | 40 (65.6%) | 148 (68.8%) | 59 (76.6%) | 22 (78.6%) | 0.352 |
| **Current BMI kg/m2 (age >18 yrs)** | 31.6 (26.1-37.5) | 28.5 (24.7-33.1) | 29.5 (24.8-33.7) | 33.4 (28.0-40.5) | 0.003 |
| **Waist:Hip Ratio** | 0.9 (0.9-1.0) | 0.9 (0.9-1.0) | 0.9 (0.9-1.0) | 0.9 (0.9-1.0) | 0.313 |
| **C-peptide (pmol/L)** | 518.0 (156.0-847.0) | 602.0 (301.0-1023.0) | 384.0 (205.0-631.0) | 354.5 (245.5-751.5) | 0.003 |
| **HbA1c (mmol/mol)** | 58.0 (45.0-77.0) | 67.0 (52.0-84.0) | 71.0 (52.0-91.0) | 72.0 (46.0-83.0) | 0.115 |
| **HDL (mmol/L)** | 1.2 (1.0-1.4) | 1.1 (0.9-1.3) | 1.3 (1.1-1.6) | 1.2 (1.0-1.5) | 0.002 |
| **Triglycerides (mmol/L)** | 1.4 (0.9-1.9) | 1.5 (1.2-2.4) | 0.9 (0.6-1.4) | 1.4 (0.8-2.0) | <0.001 |
| Data shown are n (%) or median (IQR) Median (IQR): p value by Kruskal-Wallis test Frequency (%): p value by Pearson chi2 test | | | | | |

Supplementary Table 4: Regional population data of individuals with type 2 diabetes diagnosed <30 years of age

Overall study population N = 3,290

| **Characteristics** | **Black** | **South Asian** | **White** |
| --- | --- | --- | --- |
| **N** | 612 | 1823 | 855 |
| **Female, n (%)** | 322 (52.6) | 959 (52.6) | 495 (57.9) |
| **BMI categories** |  |  |  |
| Underweight (<20 kg/m2), n (%) | 9 (1.5) | 15 (0.8) | 19 (2.2) |
| Normal (20-25 kg/m2) | 88 (14.4) | 320 (17.6) | 222 (26.0) |
| Overweight (25-30 kg/m2), n (%) | 136 (22.2) | 608 (33.4) | 174 (20.4) |
| Obese (≥30 kg/m2), n (%) | 340 (55.6) | 763 (41.9) | 394 (46.1) |
| Missing | 39 (6.4) | 117 (6.4) | 46 (5.4) |
| Mean (SD) | 33.0 (8.8) | 30.1 (6.5) | 31.3 (9.2) |
| Median (IQR) | 32.0 (26.3-37.4) | 29.1 (25.6 – 33.6) | 29.7 (23.9 – 36.9) |
| **Proportions with BMI < cut-off** |  |  |  |
| < 25 kg/m2, n (%) | 97 (15.8) | 335 (18.4) | 241 (28.2) |
| < 27 kg/m2, n (%) | 160 (26.1) | 584 (32.0) | 305 (35.7) |
| < 30 kg/m2, n (%) | 233 (38.1) | 943 (51.7) | 415 (48.5) |
| < 35 kg/m2, n (%) | 362 (59.2) | 1,359 (74.5) | 555 (64.9) |

*Taken from latest available adult BMI measurement in patient record.

Supplementary figure 1: Parental history

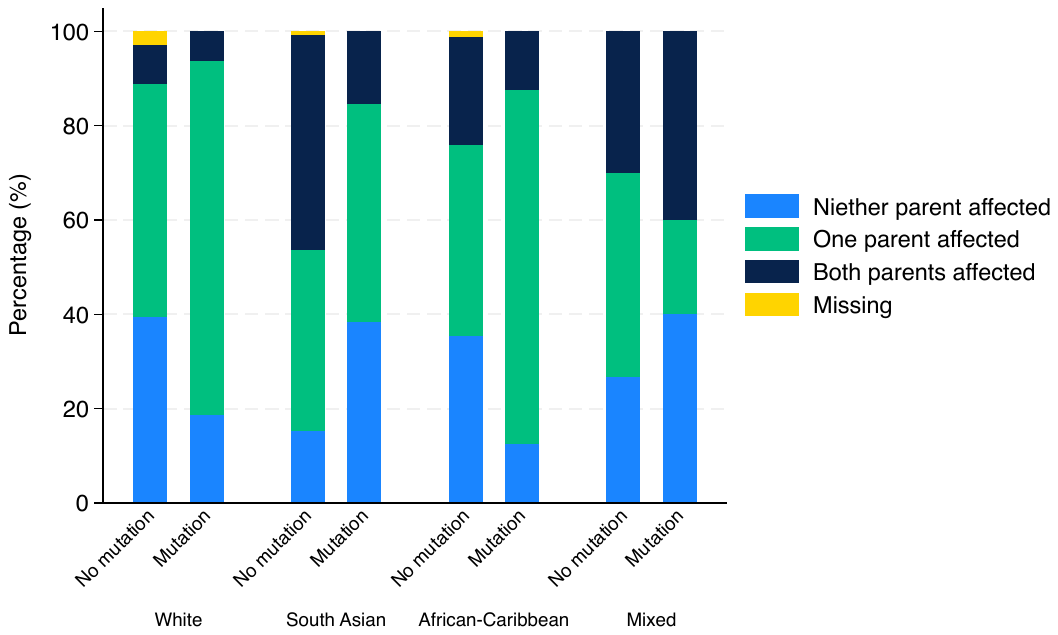

**Supplementary figure 2:** a) False negative rate and b) numbers needed to test in adult proband referrals to the National Molecular Genetics Laboratory

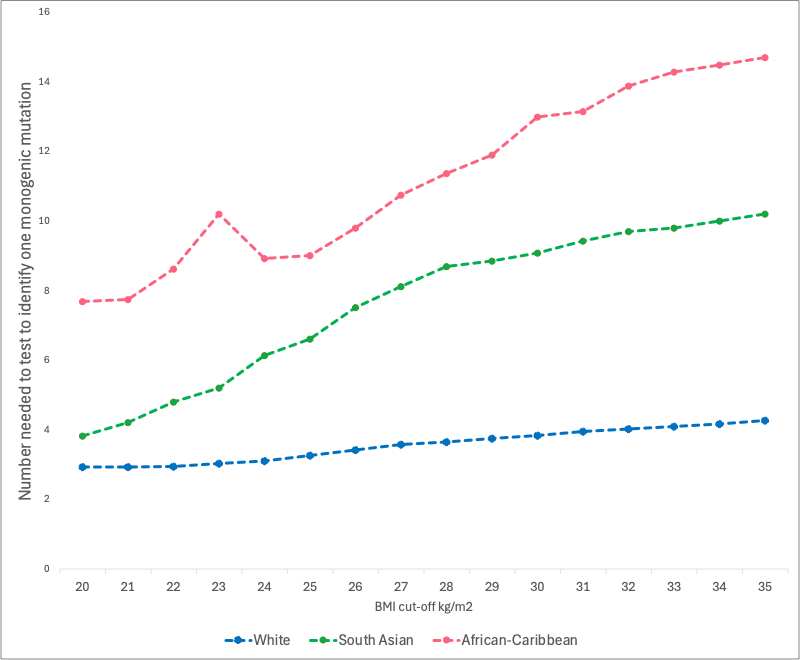

a)

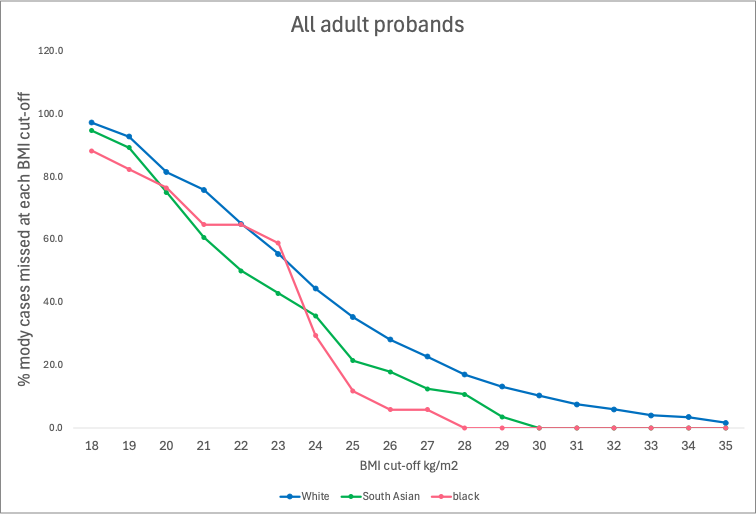

**Supplementary acknowledgements**

**List of Principal Investigators and research team members at Participating Sites**

Birmingham Children’s hospital, Dr Ruth Krone, Dr Tim Barrett & Wendy Donoghue; Birmingham Heartlands Hospital, Dr Swati Karandikar, Dr Srikanth Bellary, Laura Thrasyvoulou, Sarah Manney, Katie Price & Helen Jenner; Bradford Teaching Hospital NHS Trust: Dr Donald Whitelaw & Jo Thorpe; Bristol Royal Infirmary, Dr Natasha Thorogood and Victoria Taylor; Calderdale Royal Hospital and Huddersfield Royal Infirmary: Dr Rob Moisey, Dr Lynne Terrett and Ryan Shaw; Central Middlesex Hospital: Dr Daniel Darko and Temi Adedoyin; Chorley & South Ribble District Hospital, Dr Satyan Rajbhandari, Chandbi Sange and Wanga Ingham; University Hospital Coventry & Warickshire : Dr Heather Sterling, Dr Tom Barber, Nicola Watts, Jo Gmerek, Francesca Eardley, Gail Evans and Sue Hewins; Ealing Hospital: Dr Kevin Baynes, Sheena Quaid; Guy’s Hospital: Dr Anna Brackenridge, Olanike Okolo, Sheila Makasi; Hillingdon Hospital: Dr Giridhar Tarigopula, Natasha Mahibir, Arti Sharma; Imperial College Healthcare NHS Trust, Professor Nick Oliver; Kings College Hospital: Dr Martin Whyte and Andy Pernet; Lewisham Hospital: Dr Ruvan Kottegoda and Samia Pilgrim; Luton & Dunstable University Hospital: Dr Nisha Nathwani, Karen Samm and Karen Duncan; Manchester University NHS Foundation Trust: Dr Pushpa Jinadev, Majid Nazir, Womba Mubita and Jose Rubio; Northwick Park Hospital: Dr Mushtaq Rahman and Sean Connarty; Pinderfields Hospital: Ismaelette DelRosario and Hollie Brooke; Royal Blackburn Teaching Hospital: Dr Shenaz Ramtoola, Wendy Higginson, Jeanette Hargreaves and Claire Folan; Royal Oldham Hospital: Dr Biswa Mishra and Susan Dermody; Royal Stoke Hospital: Dr Parakkal Raffeeq, Helen Parker, Emma Sadler nd Viv Colclough; Russells Hall Hospital: Dr Haroon Siddique, Wara Mudunge and Daljit Kaur; Sandwell Children’s Hospital: Dr Chizo Agwu, Julie Oliver, Julie Colley, Carol Zullo, Julie Colley, Jenny Porter, Elizabeth Williams;

Barts Health: Dr Bobby Huda, Anne Worthington & Ken Ashida; St George’s Hospital; Dr Kenneth Earle, Vera Tavoukjian and Patricia Ribeiro; Newham Hospital, Dr Sarah Finer and Desiree Campbell-Richards; University College London Hospital: Dr Sarita Naik and Sifelani Tshuma; Walsall Manor Hospital: Dr Muhammed Javed, Liam Botfield and Robert Chadwick; Warwick Hospital: Dr Rajni Mahto, Penny Parsons and Inderjit Atwal; West Middlesex Hospital: Dr Jayanti Rangasami, Dr Rashmi Kaushal, Ursula Kirwan, Amrinder Sayan & Marie Louise Svensson; Wolverhampton Hospital: Dr Ananth Viswanath, Dr Nisha Pargass and Charlotte Busby.
